# Implementation of an Opioid Use Disorder (OUD) Machine-Learning Phenotype in Real-Time for the ADAPT Project

**DOI:** 10.64898/2025.12.02.25341524

**Authors:** Huan Li, Mark Iscoe, John Lutz, Carolina Diniz Hopper, Sabrina Fried, Josue Minaya, Caroline Raymond King, Olga Reykhart, Hyung Paek, Daniella Meeker, Edward R. Melnick, R.Andrew Taylor

## Abstract

**Objective:** Develop and deploy a real-time, EHR-integrated machine learning phenotype to identify emergency department (ED) patients with opioid use disorder (OUD) for prospective clinical trial screening and buprenorphine initiation.

**Materials and Methods:** We conducted a multi-phase study across three EDs in a single United States health system from 2014 to 2025. Using visit-level data available at or before triage, we trained a random-forest classifier to estimate OUD risk and embedded scoring in the EHR to trigger point-of-care alerts for trial eligibility review. A computable silver-standard label supported retrospective development; a clinician gold-standard reference was established via structured, DSM-5–aligned chart review. Performance was summarized with ROC-/PR-AUC, calibration, and threshold-based classification metrics; prospective validation used a stratified random sample of flagged and unflagged encounters.

**Results:** Retrospective discrimination compared to the silver standard was high (ROC-AUC 0.99, 95% CI 0.98-0.99; PR-AUC 0.92, 95% CI 0.90-0.94), and calibration plots informed operating-point selection for real-time use. In prospective gold-standard validation (n=218), the positive predictive value was 98.28%, and the negative predictive value was 95.68% at the prespecified threshold.

**Discussion and Conclusion:** An EHR-embedded, machine-learning phenotype can accurately and feasibly identify ED patients with OUD in real time, streamlining clinical trial enrollment and treatment initiation. Ongoing work will report operational metrics (e.g., alert volume and latency), monitor performance drift and equity across subgroups, and evaluate downstream clinical and trial outcomes.

**Author Summary:** In this study, we wanted to find a better way to identify people visiting the emergency department who may be living with opioid use problems. These patients are often hard to recognize quickly, even though timely support, including offering effective medications, can greatly improve their health and safety. Traditional approaches rely heavily on clinicians noticing certain patterns in real time, which can be challenging during busy emergency care. To address this gap, we examined over a decade of information from a large health system and developed a clinical decision support tool that reviews routine information collected at the start of an emergency visit. The tool gives clinicians a simple, real-time signal when a patient may benefit from additional evaluation or treatment. Our early experience shows that this approach can help teams identify patients more consistently and connect them to care more quickly. We hope this work will make it easier for hospitals to support people affected by opioid use and to run clinical studies that improve future treatment options.

## Introduction

The opioid crisis continues to evolve as one of the most urgent and complex public health emergencies in the United States. The number of overdose deaths attributed to fentanyl analogs rose by 540% from 2013 to 2016, and in 2022 alone, more than 109,000 drug overdose deaths occurred nationwide, and nearly 70% of these involved opioids.^1,2^ Emergency departments (EDs) and frontline services, including emergency medical services, are frequently the first or only point of healthcare contact for individuals with opioid use disorder (OUD) at high risk of overdose. In 2018, opioids were involved in roughly one-quarter of all drug-related ED visits across the United States, and recent studies indicate that the COVID-19 pandemic further intensified opioid-related ED utilization.^3,4^ Thus, EDs are critical access points for detection and intervention for OUD.^3,4^

Buprenorphine is a medication for opioid use disorder (MOUD) with evidence for both its effectiveness at reducing illicit opioid use and opioid-related mortality and the viability of initiating use in the ED setting.^5–8^ Despite the availability of an effective treatment and intervention, only a small subset of eligible patients, approximately 6-8%, receive buprenorphine in the ED,^9^ with limitations in its prescription and uptake related to prescribers’^5–8^ lack of experience with the medication, concerns for impact on workflow and drug diversion, and persisting stigma.^10,11^ Our group’s prior initiative, the EMBED trial, attempted to mitigate some of these gaps by developing a clinical decision support (CDS) tool for buprenorphine initiation in the ED, but was ineffective at increasing patient-level rates of buprenorphine initiation.^12^ In the ongoing ADAPT trial, we aim to build upon the work in EMBED and improve intervention effectiveness through a Multiphase Optimization Strategy Trial.^13^ A crucial step toward enhancing the adoption of ED-initiated buprenorphine is the accurate identification of potentially eligible OUD patients. Automated detection of OUD within electronic health records could enable earlier intervention, facilitate clinical trial enrollment, and strengthen linkage to treatment. Patient phenotyping through machine learning offers a practical solution to real-time patient identification.

Previous studies have used machine learning to stratify individuals according to their risk of developing or currently suffering from OUD and opioid overdose based on electronic health record (EHR) data. While several models have shown a strong predictive performance in retrospective settings,^14–16^ a lack of workflow integration has often limited their real-world application. In parallel, Chartash et al. in the EMBED trial^17^ and Taylor et al.^18^ have demonstrated that phenotypes using structured and nuanced data from patient charts can accurately identify patients with opioid-related disorders presenting to the ED. However, these phenotypes were retrospective and lacked real-time identification of eligible patients at the point of care.

To address these gaps, we developed and implemented a real time, EHR integrated, machine learning OUD phenotype to identify ED patients for prospective clinical trial screening and to support ED initiated buprenorphine as part of a nationally disseminated, multicomponent clinical decision support (CDS) program.^13^ We hypothesized that a calibrated, real time phenotype, embedded within routine ED workflow, would achieve high sensitivity with acceptable specificity at a prespecified threshold, thereby enabling timely identification of candidates for treatment and rigorous prospective evaluation.

## Results

### Participants and Descriptive Data

In the retrospective observation phase of this study, a total of 3,177,664 unique patients met the inclusion criteria, representing all adult patients with ED visits in the health system between 2014 and 2025. Following outcome-based balancing, the training dataset included 17,054 patients, and the test dataset included 4,246 patients. Within the overall population, 1,336,335 (42.05%, **Table 1**) were aged 18-39 years, 972,293 (30.60%) were aged 40-64 years, and 869,016 (27.35%) were 65 years or older. These age group distributions were similarly reflected in both the training and test datasets after balancing. In the overall population, 1,707,449 (53.73%) were female and 1,466,587 (46.15%) were male, with comparable distributions maintained in the training and test datasets. The racial/ethnic composition of the overall population was 59.81% White or Caucasian, 10.21% Black or African American, and 14.42% Hispanic. This composition was also similar in both the training and test datasets after balancing.

**Table 1.** Study Population Demographics.

|  | Overall Population<br>(n = 3,177,664) | Balanced Training Dataset<br>(n = 17,054) | Balanced Testing Dataset<br>(n = 4,246) |
| --- | --- | --- | --- |
| **Age group, n (%)** |  |  |  |
| 18-39 | 1,336,335 (42.05%) | 7,127 (41.79%) | 1,761 (41.47%) |
| 40-64 | 972,293 (30.60%) | 5,642 (33.08%) | 1,408 (33.16%) |
| 65+ | 869,016 (27.35%) | 4,285 (25.13%) | 1,077 (25.37%) |
| **Sex, n (%)** |  |  |  |
| Female | 1,707,449 (53.73%) | 8,799 (51.59%) | 2,200 (52.28%) |
| Male | 1,466,587 (46.15%) | 8,246 (48.35%) | 2,023 (47.64%) |
| Other or Unknow | 3,628 (0.11%) | 9 (0.05%) | 3 (0.07%) |
| **Race, n (%)** |  |  |  |
| White or Caucasian | 1,900,669 (59.81%) | 10,897 (63.90%) | 2,788 (65.66%) |
| Black or African American | 324,332 (10.21%) | 1,931 (11.32%) | 452 (10.65%) |
| Asian | 109,581 (3.45%) | 569 (3.34%) | 116 (2.73%) |
| American Indian or Alaska Native | 9,182 (0.29%) | 62 (0.36%) | 10 (0.24%) |
| Native Hawaiian or Other Pacific Islander | 4,962 (0.16%) | 20 (0.12%) | 10 (0.24%) |
| Other or Unknown | 828,938 (26.09%) | 3,575 (20.96%) | 870 (20.49%) |
| **Ethnicity, n (%)** |  |  |  |
| Non-Hispanic | 2,058,420 (64.78%) | 12,461 (73.07%) | 3,112 (73.29%) |
| Hispanic or Latino | 458,068 (14.42%) | 2,758 (16.17%) | 682 (16.06%) |
| Other or Unknown | 661,176 (20.81%) | 1,835 (10.76%) | 452 (10.64%) |
| **PMH, n (%) >=1** |  |  |  |
| Substance Use Disorder Diagnoses | 49,863 (1.57%) | 1,536 (9.01%) † | 379 (8.93%) † |
| Mental Diagnoses | 107,142 (3.37%) | 1,723 (10.10%) | 452 (10.65%) |
| **Chronic Illnesses** |  |  |  |
| Diabetes | 324,586 (10.21%) | 2,245 (13.16%) | 585 (13.78%) |
| Heart Disease | 280,286 (8.82%) | 2,023 (11.86%) | 526 (12.39%) |
| Hypertension | 773,361 (24.34%) | 5,286 (31.00%) | 1,335 (31.44%) |
| **Medication** |  |  |  |
| MOUD* | 58,178 (1.83%) | 1,336 (7.83%) | 375 (8.83%) |
| Other opioids | 817,469 (25.73%) | 6,081 (35.66%) | 1,547 (36.43%) |
| Naloxone | 264,999 (8.34%) | 2,624 (15.39%) | 698 (16.44%) |

\* MOUD stands medication of opioid use disorder and includes methadone, buprenorphine, naltrexone)
† Substance use disorder diagnoses defined after labeling patients with > 250 cumulative OUD codes as positive.

In terms of relevant past medical history (PMH), 1.57% of the overall population had a documented substance use disorder, 3.37% had mental health diagnoses, and among selected chronic illnesses, 10.21% had diabetes, 8.82% had heart disease, and 24.34% had hypertension. Regarding medication use, 1.83% had prescriptions for medications for opioid use disorder (MOUD: methadone, buprenorphine, or naltrexone), 25.73% for other opioids, and 8.34% for naloxone.

Unlike demographic characteristics, PMH and medication prevalence, by design, differed between the training and test datasets due to the balancing procedure. In the balanced training set, prevalence rates were higher: 9.01% for substance use disorder, 10.10% for mental health diagnoses, 13.16% for diabetes, 11.86% for heart disease, and 31.00% for hypertension. Corresponding rates in the test set were similar: 8.93%, 10.65%, 13.78%, 12.39%, and 31.44%, respectively. MOUD use was 7.83% in training and 8.83% in testing; other opioid use was 35.66% and 36.43%; naloxone med list rates were 15.39% and 16.44%.

### Phenotype Model Outcomes

The random forest classifier model predicted the patients with a positive OUD phenotype (based on the sliver standard) with a sensitivity of 0.549, specificity of 0.997, precision of 0.945, NPV of 0.958, FPR of 0.003, FNR of 0.451, accuracy of 0.957, and F1 score of 0.694, when evaluated at the default probability threshold of 0.50. Additional evaluations using alternative probability thresholds are presented in **Table 2**.

**Table 2.**
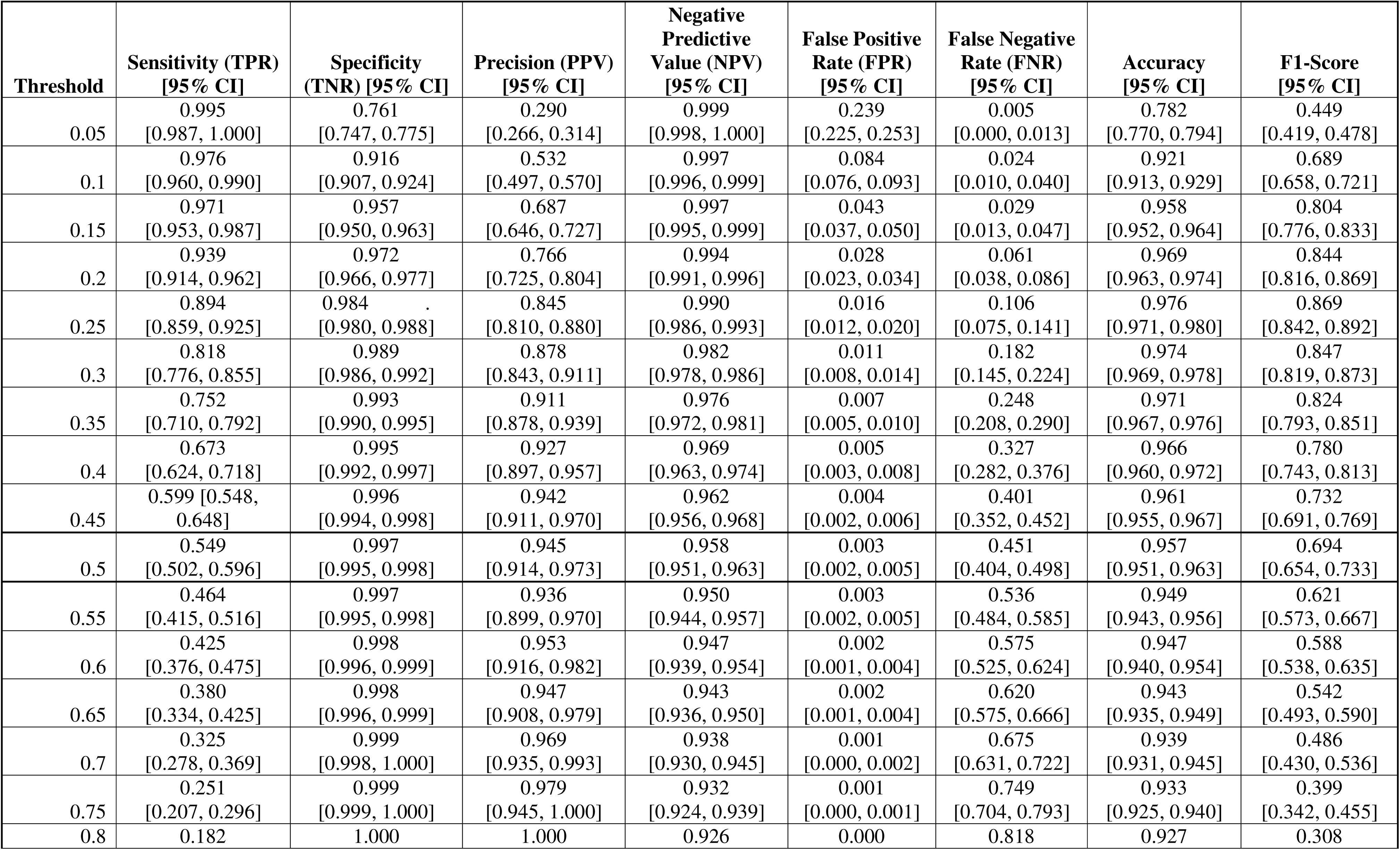
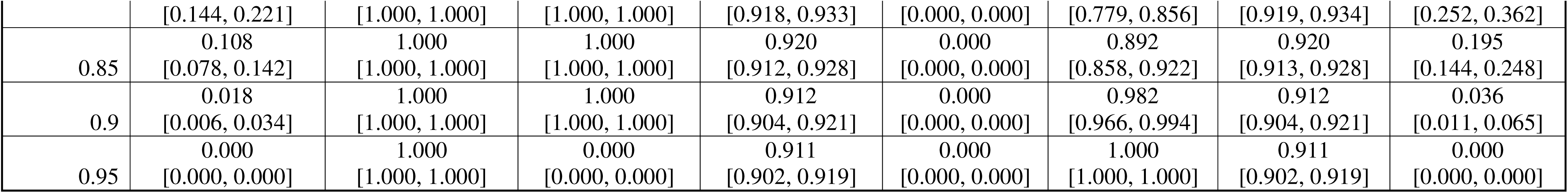
Performance Metrics of the Predictive Model.

At the default probability threshold of 0.50, the confusion matrix demonstrated 202 true positives, 10 false positives, 3805 true negatives, and 172 false negatives (**Figure 1a**). The ROC curve yielded an AUC of 0.99 (95% CI 0.98-0.99), indicating strong overall discrimination between patients with and without OUD (**Figure 1b**). The precision–recall curve produced an AUC of 0.92 (95%CI 0.90-0.94), supporting the model’s ability to preserve positive predictive value under class imbalance (**Figure 1c**). Finally, the calibration curve demonstrated that predicted probabilities were underestimated across the observed risk spectrum, especially for higher predicted probabilities (**Figure 1d**).

**Figure 1.**
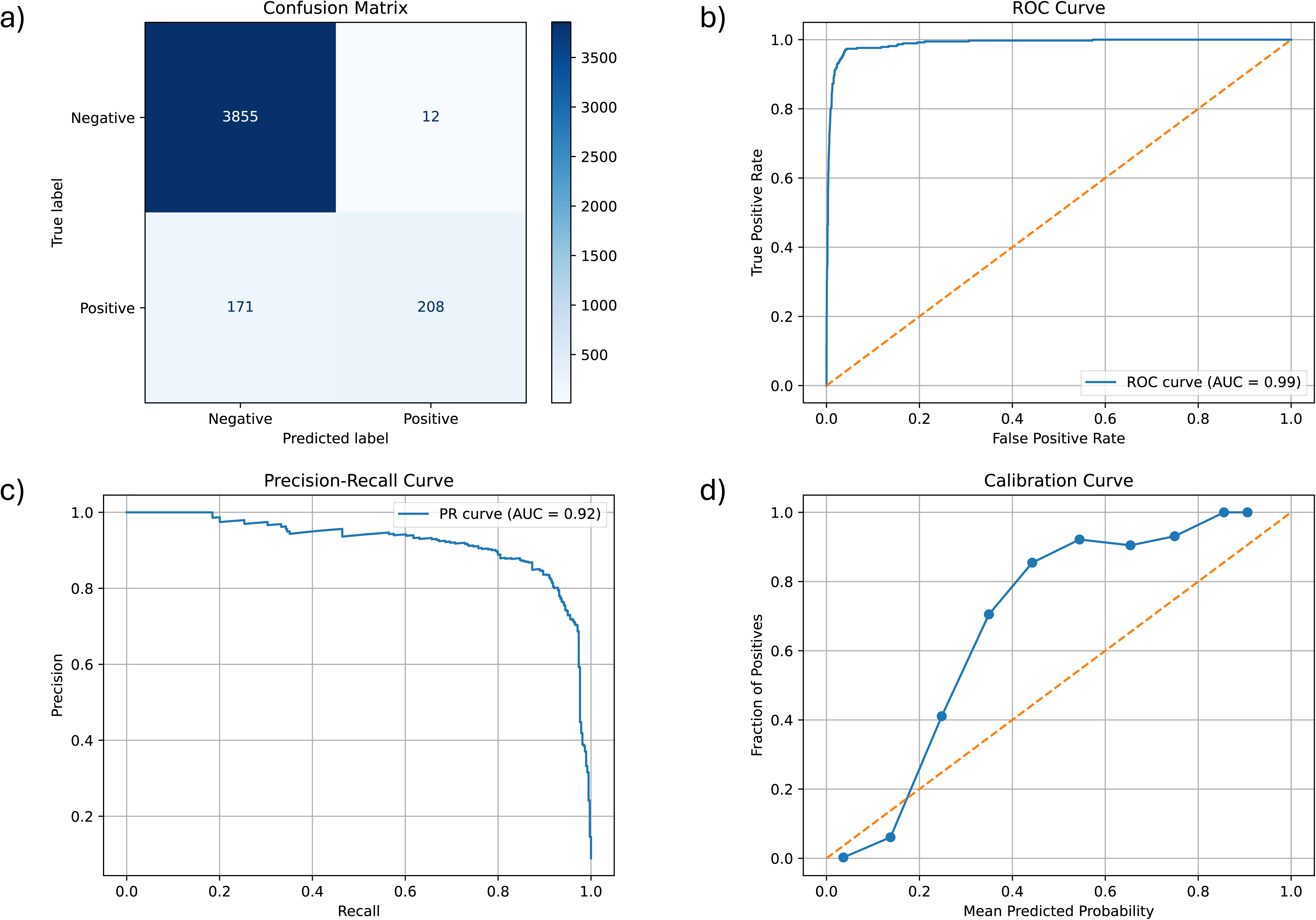
Silver Standard Prediction Model Performance Evaluations: a) confusion matrix at the default probability threshold of 0.50, b) Receiver operating characteristic (ROC) curve with area under the curve (AUC), c) precision-recall (PR) curve with area under the curve, d) calibration curve comparing predicted versus observed probabilities.

The top 25 predictive features, ranked by feature importance, derived from the random forest classifier, are presented in **Figure 2** (full list in **Appendix 1**). At the individual feature level, the most influential predictors of the OUD phenotype were diagnoses related to substance use disorders other than OUD. At the category level, features pertaining to PMH contributed the greatest overall importance, followed by medication-related variables and laboratory results. Notably, the top two of the three individual features belonged to the PMH category.

**Figure 2.**
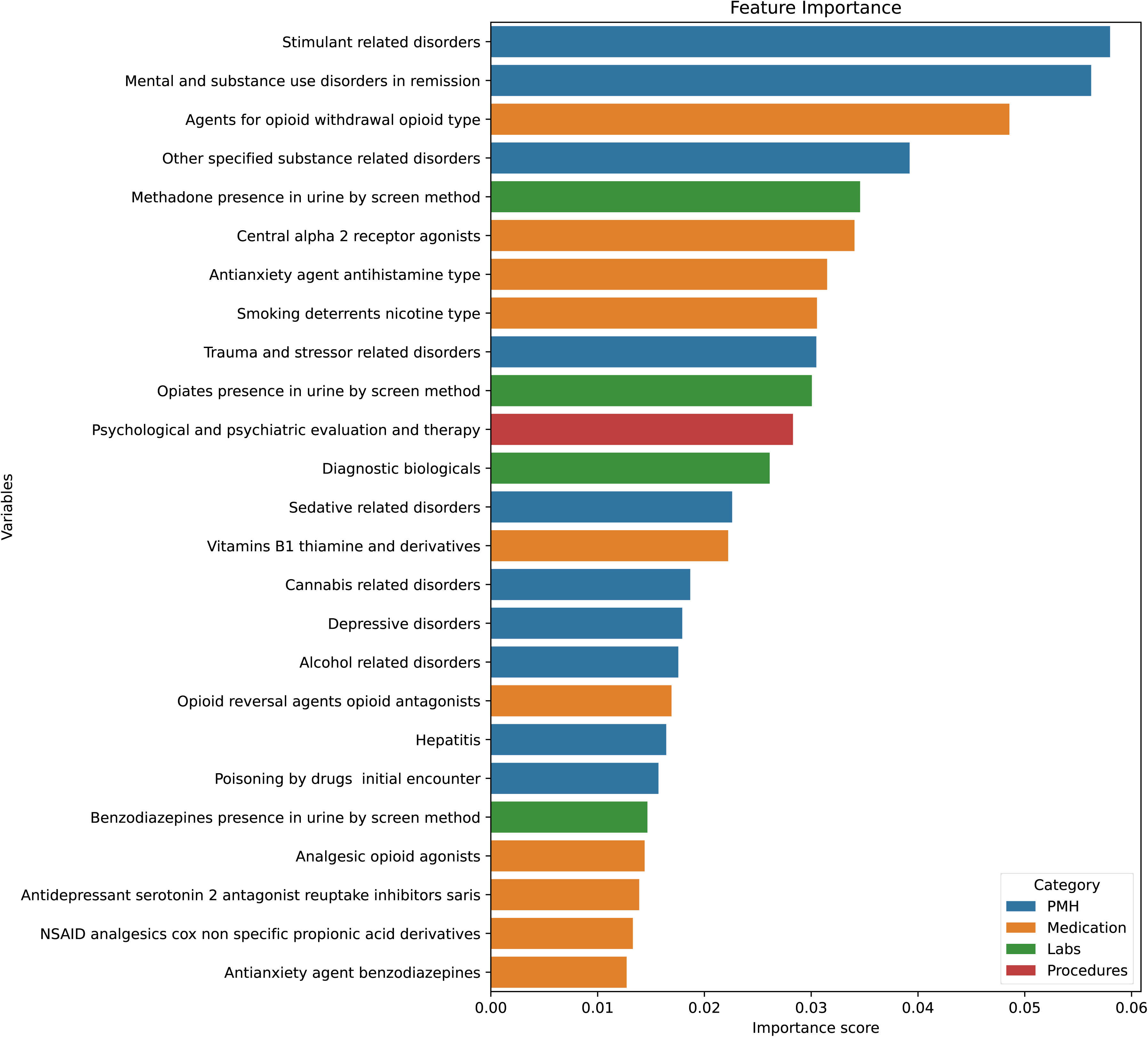
Feature Importance of Predictors for OUD Phenotype: Bar size represents the relative importance of each of the top 25 predictors, as estimated by the random forest classifier. Colors indicate feature categories: PMH (blue), medication (orange), labs (green), procedures (red), and other (purple).

### Gold Standard Evaluation

In the prospective validation phase of the study, there were 217 patients in the gold standard validation set, including 96 (44.24%, **Table 3**) classified by the model as having OUD and 121(55.76%) classified as not having OUD. We initially selected 200 patients for chart review and subsequently added 20 additional cases (for a total of 220) to ensure sufficient sample size after applying exclusion criteria. Three patients were excluded (one younger than 18 years and two who were pregnant), resulting in the final cohort size reported above. Of these patients, 42.86% (n = 93) were female and 57.14% (n = 125) were male. The age distribution was as follows: 24.54% (n = 53) were aged 18–39 years, 48.15% (n = 105) were aged 40–64 years, and 27.31% (n = 60) were aged 65 years and older. Regarding race and ethnicity, 67.28% (n = 147) of patients identified as White or Caucasian, 22.58% (n = 49) as Black or African American, 19.32% (n = 43) as Hispanic or Latino, and 0.46% (n = 1) as Asian. After chart review, 103 were ultimately adjudicated as having OUD and 114 as not having OUD. Cohen’s kappa for inter reviewer agreement was 0.86. Of the 17 cases in which resident reviewers disagreed on patients’ OUD status, attending physician adjudicators initially agreed on 13 (76%) and reached a consensus on the remaining 4 cases after discussion. The demographic composition of the validation set was broadly comparable between patients with and without OUD, as determined by gold standard manual chart review. The age distribution differed slightly, with a higher proportion of patients aged 40–64 years in the OUD positive group and a higher proportion aged 65 years and older in the OUD negative group.

**Table 3.** Gold Standard Validation Group Demographics.

|  | Overall (n = 217) | OUD Yes (n = 103) | OUD No (n = 114) |
| --- | --- | --- | --- |
| Age group, n (%) |  |  |  |
| 18-39 | 53 (24.54%) | 26 (25.24%) | 27 (23.89%) |
| 40-64 | 104 (48.15%) | 60 (58.25%) | 44 (38.94%) |
| 65+ | 59 (27.31%) | 17 (16.50%) | 42 (37.17%) |
| **Sex, n (%)** |  |  |  |
| Female | 93 (42.86%) | 39 (37.86%) | 54 (47.37%) |
| Male | 124 (57.14%) | 64 (62.14%) | 60 (52.63%) |
| Unknow | 0 |  |  |
| **Race, n (%)** |  |  |  |
| White or Caucasian | 146 (67.28%) | 75 (72.82%) | 71 (62.28%) |
| Black or African American | 49 (22.58%) | 20 (19.42%) | 29 (25.44%) |
| Asian | 1 (0.46%) | 0 | 1 (0.88%) |
| American Indian or Alaska Native | 0 | 0 | 0 |
| Native Hawaiian or Other Pacific Islander | 0 | 0 | 0 |
| Other or Unknown | 22 (10.14%) | 8 (7.77%) | 13 (11.40%) |
| **Ethnicity, n (%)** |  |  |  |
| Non-Hispanic | 170 (78.34%) | 81 (78.64%) | 89 (78.07%) |
| Hispanic or Latino | 43 (19.82%) | 21 (20.39) | 22 (19.30%) |
| Unknown | 4 (1.84%) | 1 (0.97%) | 3 (2.63%) |

Model performance evaluated against the gold standard OUD labels in the validation set (**Figure 3**), identified 92 true positives (TP), 110 true negatives (TN), 11 false positives (FP), and 5 false negatives (FN, **Figure 3**). The model yielded a sensitivity of 0.958 (95% CI: 0.898 - 0.984) and a specificity of 0.909 (95% CI: 0.845 - 0.948) with respect to the gold standard.

**Figure 3.**
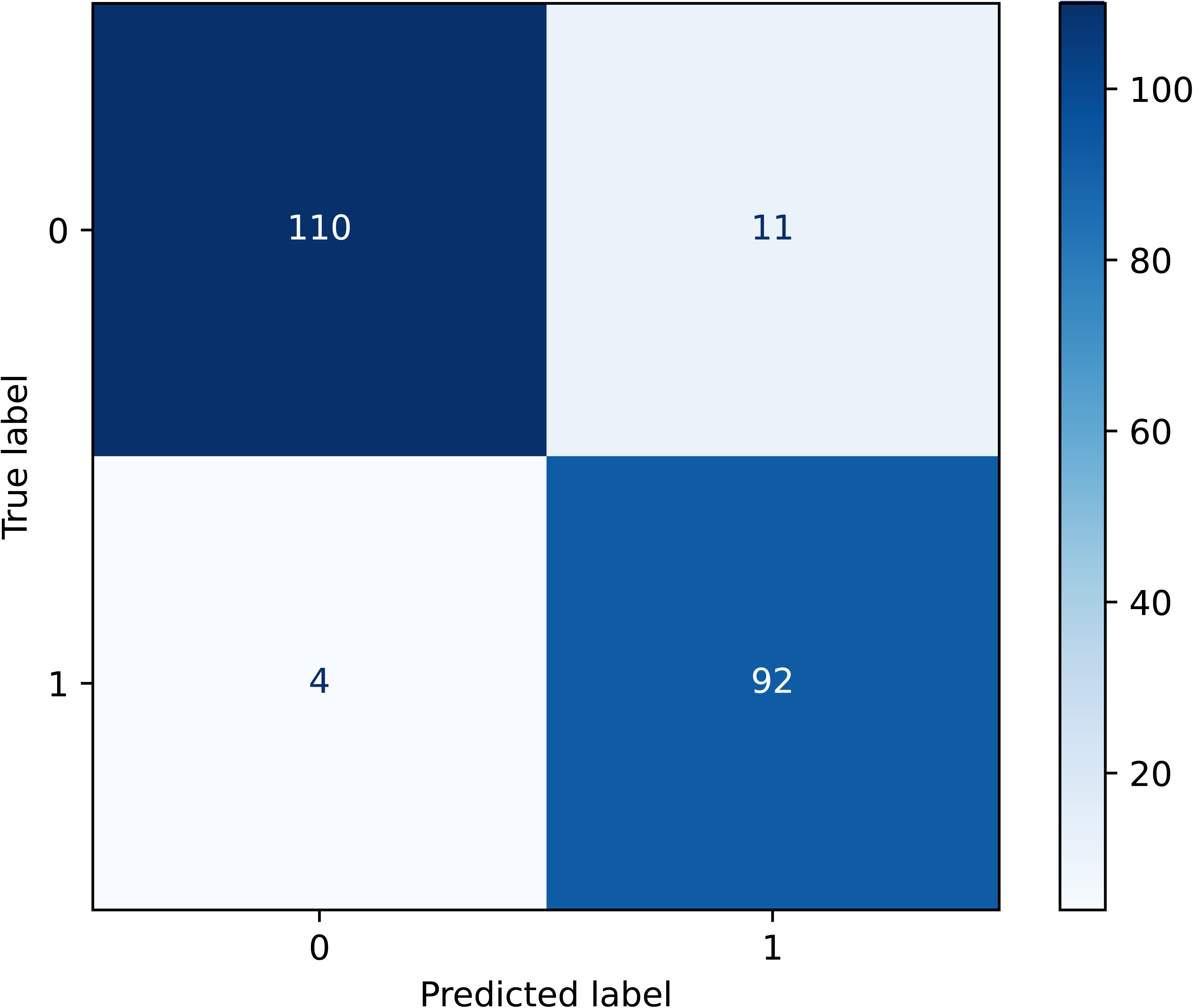
Gold Standard Validation Outcome Confusion Metrics. The figure presents the distribution of TP, FP, TN, and FN from the validation set. Each cell represents the count of patients correctly or incorrectly classified by the model after chart review based on the gold standard. The model demonstrated high agreement with the gold standard.

## Discussion

In this multi-phase study, we developed, implemented, and prospectively validated an EHR embedded phenotype capable of accurately identifying patients with OUD in real time. By integrating daily inference within existing data infrastructure, we demonstrated that automated phenotyping can transition from retrospective research to prospective clinical application with minimal latency. The model’s high predictive performance and operational feasibility highlight its potential to enhance clinical care and clinical trial efficiency for ED patients with OUD.

The phenotype classifier model demonstrated excellent performance in identifying patients with OUD, achieving a high AUC of 0.99 in the silver standard evaluation based on diagnostic codes, and a sensitivity of 0.96 and specificity of 0.91 in the gold-standard validation based on physician adjudication of charts; this performance is substantially higher than values reported in prior studies using EHR-based prediction models for substance use disorders, which have ranged from 0.80 to 0.95 with the majority falling in the 0.85 - 0.90 range.^19,20^ The strong performance observed in both retrospective and prospective validation phases highlights the model’s robustness and reproducibility. Several factors may have contributed to this strong performance, including the use of a high-quality outcome definition (silver standard), the construction of features from detailed EHR data that captured robust patterns indicative of OUD, and the application of tree-based models, which are well-suited for handling non-linear relationships and complex interactions among clinical features. Additionally, the model’s predictions were highly consistent with the gold-standard chart review outcomes, demonstrating strong alignment between algorithmic classification and clinical validation.

The pipeline was designed to operate within standard EHR data structures and executed efficiently using distributed computing, demonstrating technical feasibility for real-time integration. Our implementation also differs from prior work by closing the loop operationally— nightly OMOP harmonization, daily inference, and EHR-native rules—yielding ≤ 24-hour latency.^21^ These findings suggest that real-time deployment within the EHR environment is both technically feasible and clinically effective, enabling timely identification of patients at risk for OUD who could benefit from appropriate ED intervention and treatment. While the EMBED pragmatic trial demonstrated that CDS alone does not guarantee population-level uptake;^22^ the new phenotype presented here could address these challenges by offering a maintainable pathway for automated, real-time screening to support increased patient identification and subsequent treatment. Finally, multi-site rule sets have demonstrated portability across health systems; by grounding our pipeline in standard vocabularies and OMOP, we similarly position the phenotype for reuse and external validation.^17^

Prior ED phenotyping efforts have primarily relied on rule-based algorithms anchored to diagnosis codes and a small set of structured signals. While precise, these approaches: 1) under detect uncoded OUD, and 2) do not allow for detection and intervention during the encounter itself (given diagnostic codes are typically applied at the end of or after the encounter.^14,17,23,24^ More recent machine-learning models for OUD and overdose risk report strong retrospective discrimination, but most remain decoupled from real-time, clinical workflow, with limited attention to calibration, latency, or operational usability.^18^ In contrast, our strategy paired a high-specificity silver standard for model development with DSM-5–aligned gold-standard adjudication for prospective evaluation. This approach reduced label noise by limiting misclassified or ambiguous cases in the training data, while quantifying real-world performance. Unlike diagnostic code-only phenotypes, we used count-based longitudinal features (diagnoses, medications, laboratory positives) to preserve burden and utilization signal, which may explain the higher discrimination observed. Finally, few prior studies report fairness or drift monitoring plans;^14,17^ we explicitly outline future equity audits and recalibration, which are essential for sustained, trustworthy deployment in dynamic ED environments.

Our findings demonstrate that an electronic phenotype can reliably identify patients with OUD in the ED in real time, offering a practical bridge between predictive modeling and actionable clinical use. Such integration has immediate implications for both research and care delivery: it can streamline enrollment in clinical trials such as the ADAPT Trial,^13^ reducing manual screening burden and ensuring that eligible patients are recognized and offered evidence-based treatments like buprenorphine. Beyond trial operations, real-time OUD detection can facilitate more consistent linkage to addiction medicine and recovery resources, reducing variability across clinicians and encounters. As health systems expand the use of phenotyping pipelines, this approach could serve as a generalizable model for identifying other high-risk conditions requiring timely intervention in the ED and beyond.

This study has several limitations. First, it was conducted within a single academic health system using a single EHR vendor product and locally harmonized data; results may not generalize to other institutions, vendors, or patient populations. Second, the retrospective “silver-standard” label, based on high counts of OUD-related ICD codes, may misclassify patients due to documentation or utilization bias. Third, the prospective validation used stratified sampling based on model predictions rather than natural prevalence, so we were not able to directly report sensitivity and specificity, limiting external validity of performance values. Fourth, the model relied on structured EHR data, omitting the potentially-informative narrative content of clinical notes, and used simple imputation method (mode) that may introduce bias by over representing common values and obscuring true variability across patients or sites; undersampling and within system validation may also overestimate performance. Finally, although demographic variables were excluded from modeling, bias may persist through proxy features.

Future work should focus on evaluating the clinical and operational impact of real-time OUD phenotyping in prospective, multi-site trials. Specifically, studies should assess whether automated identification improves rates of ED-initiated buprenorphine, treatment linkage, and long-term recovery outcomes. Ongoing monitoring for performance drift, calibration decay, and subgroup disparities will be critical to maintaining accuracy and fairness over time. Finally, extending this framework to other conditions, such as alcohol use disorder, suicidality, or sepsis in the ED (or beyond) could demonstrate the broader potential of embedded phenotyping to enhance real-time clinical decision support.

## Materials and Methods

### Study Design and Study Population

This multi-phase study involved two components: 1) a retrospective observational study utilizing ED visit data from 2014 to 2025 to develop a predictive model, and 2) a prospective validation of the resulting model. The validated model was created with the goal of integration into a clinical decision support tool designed to identify OUD patients eligible for buprenorphine initiation in the prospective ADAPT clinical trial.^13^ This manuscript describes model development and prospective phenotype validation in preparation for the ADAPT trial; results of the trial itself will be reported after trial completion.

The model was trained on patient data from across the regional health system, including nine EDs. The study population for this phase included all adult patients aged 18 and older who had not opted out of research use. This study employed the STROBE reporting guidelines for observational research.^25^ All data processing, feature engineering, and model development were performed using PySpark (*PySpark v3.2.1*) on our health system’s local cloud-based secure computing platform. The descriptive statistics of the study population, including the overall cohort, the train/test subsets (after balancing and splitting), are summarized in **Table 1**. Phenotype development was deemed exempt from human subject review by our institution’s IRB (protocol # 2000037541), and prospective use in the ADAPT clinical trial was approved (protocol # 2000038624) with a waiver of informed consent. The ADAPT clinical trial is registered on clinicaltrials.gov (ID: NCT06799117).

### Data Source, Variables, and Measurements

All patient data were extracted from the health system’s Epic EHR (Epic, Verona, WI) data systems (Clarity and Caboodle, **Figure 4**, pipeline diagram). Using established extract– transform–load (ETL) procedures, records were ingested into the local Computational Health Platform (CHP) and harmonized to the OMOP common data model for analysis. Routine data quality checks (completeness, conformance, and plausibility) were applied at each stage of the pipeline. Demographic information, including age, gender, race, and ethnicity, was collected but not included as model features. Past medical history items were categorized according to Agency for Healthcare Research and Quality (AHRQ) Clinical Classification Software (CSS) categories,^26^ along with an additional variable that counted instances of OUD based on the provided ICD code value set (**Appendix 2**).^27^ Medication exposures and orders were summarized as counts rather than simple presence/absence, and for pre-specified laboratory tests, we recorded the number of positive results. All feature tables were joined on the unique patient identifier. Then, variable names were normalized (lowercase; special characters removed), and missing values were imputed using the variable-wise mode. This count-based representation was selected because it preserves longitudinal burden and utilization signal compared with binary indicators.

**Figure 4.**
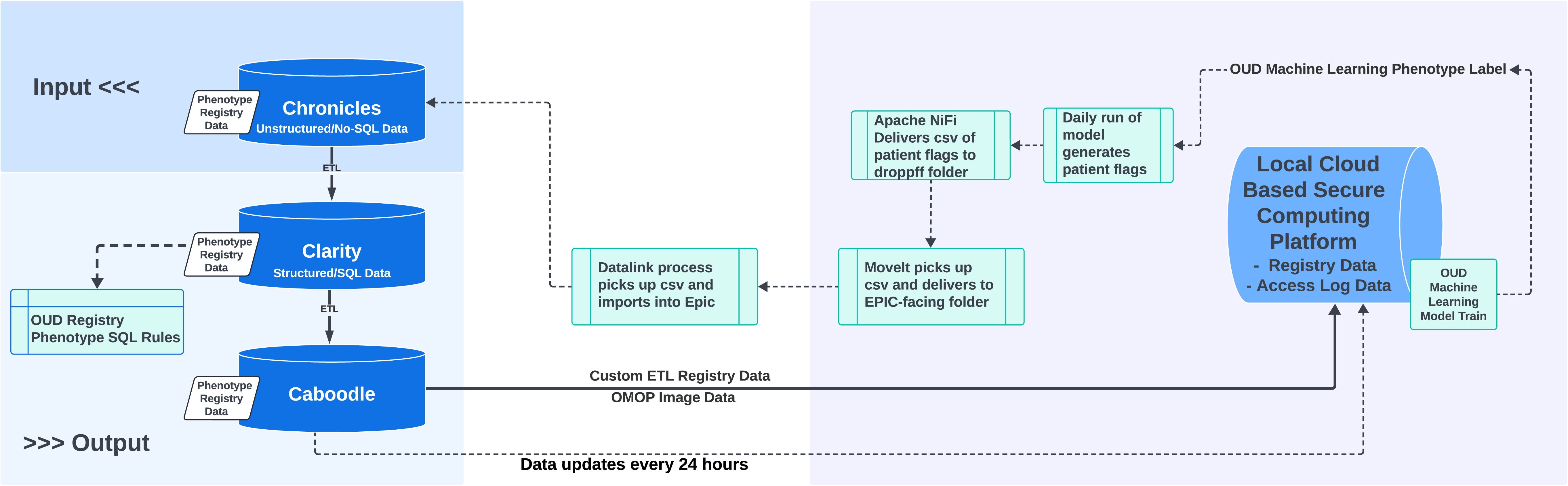
System Architecture and Data Flow: This pipeline diagram illustrates the multi-phase process of data abstraction, model development, and deployment of a pre-trained OUD prediction model. It displays how the model was integrated into the EHR system to support real-time CDS for the timely identification of eligible ED patients. Chronicles is Epic’s foundational operational database that stores real-time patient-level clinical data. Clarity is a relational reporting database that extracts and standardizes data from Chronicles for analytical use. Carboodle is an analytics-ready data warehouse built on top of both Chronicles and Clarity, designed to support research and operational reporting. The local cloud-based secure computing platform serves as the environment where the model is built, validated, and operated, using the OMOP common data model, which is a Common Data Model that YNHH database engineers extract from the Epic Caboodle database according to OHDSI specifications. Apache NiFi, deployed via Docker, facilitates automated and reproducible data ingestion and transformation workflows across systems. Within Epic, Datalink was utilized to enable targeted queries and cohort selection directly from the EHR.

#### Silver Standard

Since OUD prevalence in our ED cohort is low and manual physician adjudication is resource-intensive, we trained the model using a silver-standard proxy rather than a physician-adjudicated gold standard. The silver-standard outcome was defined from longitudinal EHR data as the cumulative count of OUD-related ICD codes (value set in **Appendix 2**) recorded in each patient’s medical record. Patients with >250 cumulative OUD codes were labeled positive for OUD. This high threshold was selected to avoid false positives from high-frequency copy-and-paste documentation habits as well as diagnosis codes existing in multiple places in the chart (such as problem list, past medical history, and encounter diagnoses), resulting in a bloated number of diagnosis codes and refined through targeted clinical chart reviews to prioritize specificity and maximize positive predictive value (PPV)^28^, while also optimizing sensitivity (**Appendix 3**). For downstream flagging, we created a composite operational label to improve sensitivity: a patient was considered positive if either (i) the model predicted OUD or (ii) the cumulative OUD ICD count was ≥25. This rule preserved the high specificity anchor used for training while broadening capture to patients with substantial—but less extreme—documentation density.

#### Gold Standard

We used physician chart review consistent with the Diagnostic and Statistical Manual of Mental Disorders, Fifth Edition (DSM-5) definition of OUD as our gold standard,^29^ with full methods detailed provided in **Appendix 4**.

## Model Development and Validation

### Phenotyping Prediction Algorithm

A random forest classifier was utilized to predict patient OUD status at the start of an ED encounter based on the silver standard definition (XGBoost was not available in our PySpark, secure compute environment; we also evaluated logistic regression (with and without L1/LASSO)), but Random Forest delivered higher AUROC/PR-AUC and comparable calibration on cross-validation, so it was selected as the primary model). Tree-based models are well known for their strong generalization performance, high predictive accuracy, and interpretability.^30^ To address the imbalance in the dataset, we applied undersampling to the majority class (non-OUD) through random sampling based on sliver standard defined outcomes, thereby aligning its frequency with that of the positive class (OUD), and achieved a 10:1 ratio. Subsequently, we performed feature engineering using PySpark’s VectorAssembler to consolidate relevant features into a single vector column. The balanced dataset was then divided into training and testing subsets, with an 80:20 ratio, consistent with standard practice in model development. The Random Forest classifier was trained on the training data, with hyperparameters optimized using a Binary Classification Evaluator. ^31,32^ This evaluator assessed model performance based on the area under the ROC curve (AUC), and 2-fold cross-validation was performed to mitigate the risk of overfitting.

### Evaluation Analysis

#### Phase 1 Silver Standard Evaluation

Model performance compared to the silver standard was evaluated using confusion matrices (i.e., 2×2 contingency tables, **Figure 1a**), along with additional metrics including sensitivity (recall), specificity, PPV (precision, the proportion of predicted positives that were true positives), negative predictive value (NPV), false positive rate (FPR), false negative rate (FNR), accuracy, and F1 score (harmonic mean of precision and recall, **Table 2**). The ROC AUC and the area under the precision-recall curve (PR AUC) were also calculated and plotted (**Figure 1b, c**). In addition, a calibration curve (**Figure 1d**) was plotted to assess the model’s goodness of fit. Feature importance was also generated and plotted to better understand the association between features and the outcome variable.

#### Phase 2 Gold Standard Evaluation: Chart Review

To prospectively evaluate model performance, four resident physicians (SF, JM, CRK, OR) with extensive experience caring for OUD patients conducted manual chart review of randomly selected cases. We selected cases based on model classification to address class imbalance (patients classified as not having OUD far outnumbered those classified as having OUD), randomly selecting 110 patients from each prediction class (OUD positive or negative by model prediction), with the goal of retaining at least 100 patients in each class and a total of 200 patients after exclusions. Sampling was restricted to a single visit per patient. To ensure consistency with the Epic CDS alert algorithm logic and the inclusion criteria used in prior phases, we applied exclusion criteria to remove cases involving pregnant patients or patients under 18 years of age.

Each case was reviewed by two resident physicians, who were given a medical record number and date of encounter (the date at which the model prediction was made) and asked to determine whether the patient met criteria for a diagnosis of OUD according to the DSM-5 definition at the start of the encounter; reviewers were blinded to the model’s predictions. To ensure consistency and accuracy in labeling, reviewers followed structured chart review guidance, adapted from Pulumbo et al.,^33^ as detailed in **Appendix 4**. In addition to structured data elements, reviewers were directed to review unstructured clinical notes, which could contain data and details not included in predictive model features, all of which were based on structured data elements. If reviewers noted that they had cared for a patient whose chart they were reviewing, the patient was excluded to avoid bias related to personal clinical experience.

Reviewers’ assessments of each patient’s OUD status were compared, and inter-reviewer agreement was assessed with Cohen’s kappa statistic. In cases in which both reviewers agreed on the patient’s OUD status, their assessment was considered the gold standard. In cases of disagreement, two attending physicians (EM and MI) with expertise in OUD reviewed the cases independently; if they both agreed, their assessment was considered the gold standard. If they disagreed, they discussed the case to reach a final consensus gold standard diagnosis.

Model performance with respect to the gold standard was reported as positive predictive value (PPV, the proportion of phenotype-positive cases that met gold standard criteria for OUD) and negative predictive value (NPV, the proportion of phenotype-negative cases that did not meet gold standard criteria for OUD). These metrics were chosen to directly assess the clinical relevance of the phenotype in identifying true positive and negative cases of OUD, particularly in the context of DSM-5 criteria.^17^

## Real-time Implementation

In our real-time implementation of this machine-learning OUD phenotype, on a nightly (∼24 hour) schedule, encounters and longitudinal EHR elements were extracted from Epic Clarity/Caboodle into the institutional Computational Health Platform (CHP) and harmonized to the OMOP common data model. The trained OUD phenotype model was then executed as a daily batch job across the OMOP tables to produce a patient-level phenotype probability and binary flag for all eligible patients. Flags were published back to Epic via Apache NiFi and Epic DataLink as CSV payloads to a monitored drop location and ingested into production, yielding ≤24-hour latency between documentation and flag availability (**Figure 4**).

At ED arrival/registration, the real-time phenotype rules implemented in the Epic EHR consumed the latest model flag and applied additional clinical logic. Adults (≥18 years) triggered an alert if they either met the model’s operating threshold (selected during validation to balance PPV and sensitivity) or presented with OUD-relevant chief complaints, received pre-hospital naloxone, or had triage documentation of a clinical opiate withdrawal scale (COWS) score > 7. Alerts were suppressed for patients with an existing prescription in the predefined medication grouping (given the trial’s goal to initiate medication treatment for OUD, **Appendix 5**), pregnancy, or documented research opt-out.

## Data Availability

The clinical data used in this study are derived from the electronic health record system of the Yale New Haven Health System and contain protected health information. Due to legal and ethical restrictions under HIPAA and institutional policy, these data cannot be made publicly available.

## Acknowledgment

We acknowledge the use of ChatGPT (OpenAI, GPT-5.0) and Yale’s Clarity GPT4 for facilitating advanced language modeling and Grammarly for enhancing our manuscript’s clarity and quality.

## Notes

**Funding:** Research reported in this publication was supported by the National Institute On Drug Abuse of the National Institutes of Health under Award Number R33DA059884. The content is solely the responsibility of the authors and does not necessarily represent the official views of the National Institutes of Health.

### Competing Interest Statement

The authors have declared no competing interest.

### Funding Statement

Research reported in this publication was supported by the National Institute On Drug Abuse of the National Institutes of Health under Award Number R33DA059884. The content is solely the responsibility of the authors and does not necessarily represent the official views of the National Institutes of Health.

### Author Declarations

The development of the machine-learning phenotype was determined to be exempt from human subjects review by the Yale University Human Research Protection Program Institutional Review Board (IRB Protocol #: 2000037541).

### Summary of Updates

This revision updates the metadata to correct a submission error. The manuscript PDF remains unchanged.

